# Quantitative Cerebrovascular Analysis for Improved Prediction of Post-Stroke Complications

**DOI:** 10.64898/2026.02.12.26346217

**Authors:** Aditi Deshpande, Jing Wang, Laith R Altaweel, Seajin Yi, Zelalem Bahiru, Tahddeus J Leiphart, Pouya Tahsili-Fahadan, Kaveh Laksari

## Abstract

**Background:** Endovascular thrombectomy (EVT) has transformed the treatment of acute ischemic stroke (AIS). However, a substantial proportion of AIS patients experience poor outcomes despite successful recanalization, often due to severe neurological deterioration or life-threatening complications. Early identification of these high-risk patients remains a major unmet need. In this study, we developed and validated machine-learning (ML) models that integrate automated quantitative cerebrovascular morphology and collateral grading with demographic, clinical, laboratory, and imaging variables to predict major post-EVT complications and early neurological outcomes.

**Methods:** Using a prospectively collected database of 727 AIS patients that underwent EVT, we developed ML models to incorporate patient-specific vascular morphometry with conventional clinical, laboratory, and imaging data to predict emergence of early neurological deterioration (END), symptomatic intracranial hemorrhage (sICH), malignant brain edema (MBE) requiring surgical decompression, and neurogenic respiratory failure and dysphagia requiring tracheostomy/gastrostomy (TC/PEG).

**Results:** Our analysis of morphological features, including increased tortuosity and reduced vessel diameter, showed strong associations with complications. Morphology-informed (MI) models consistently outperformed baseline-clinical (BC) models for patients with END (AUROC 0.81 for MI model vs. 0.73 for BC), sICH (AUROC 0.68 MI vs. 0.56 BC model), MBE (AUROC 0.67 MI model vs. 0.56 BC), or patients who underwent TC/PEG (AUROC 0.66MI vs. 0.58 BC model). Statistical testing confirmed significant AUROC improvements for END, sICH and mRS (p < 0.05), Finally, patient-specific calibrated probability profiles enabled individualized, multidimensional risk stratification, revealing distinct complication-specific risk patterns across patients.

**Conclusions:** These findings demonstrate that cerebrovascular structure—an often overlooked yet physiologically fundamental determinant of ischemic injury and reperfusion dynamics—provides significant predictive information that is not captured by standard clinical or visual imaging assessments. Automated vascular segmentation and collateral grading techniques enable rapid and objective integration of cerebrovascular metrics into prognostic models, offering a scalable tool for precision risk stratification, supporting earlier intervention, targeted monitoring, and improved post-EVT management.

## 1. INTRODUCTION

Acute ischemic stroke (AIS) remains a leading cause of morbidity and mortality worldwide ^1^. Acute reperfusion therapies—intravenous thrombolysis (IVT) and endovascular thrombectomy (EVT)— restore cerebral blood flow (CBF), improve long-term functional outcomes, and reduce mortality ^2^. In particular, EVT has transformed AIS management in patients with large and medium vessel occlusion. However, despite technically successful recanalization (assessed by the modified Thrombolysis in Cerebral Infarction [mTICI] score), 30–40% of AIS patients continue to experience significant morbidity after EVT, with many developing severe complications that adversely impact recovery ^3,4^. Approximately 13-29% of morbidity and mortality after stroke is directly attributed to severe, acute post-AIS complications ^5,6^.

Residual hypoperfusion, microvascular embolization, and reperfusion injury are recognized contributors to the disconnect between successful angiographic recanalization and favorable clinical outcomes ^4,7^. These mechanisms drive early neurological deterioration (END) and complications such as hemorrhagic transformation of infarcted tissue, leading to symptomatic intracranial hemorrhage (sICH), and malignant brain edema (MBE) requiring decompressive hemicraniectomy (DHC) ^8,9^. The early deterioration and complications lead to worse long-term outcomes and increased mortality^4,5^. Therefore, early and accurate prediction of these complications is critical for optimizing post-EVT management, informing decisions regarding monitoring intensity, resource allocation, and potential surgical intervention.

Multiple scoring systems have been developed to predict early post-stroke complications ^10–12^. Recent efforts using machine learning (ML) have expanded predictive modeling by incorporating various demographic, clinical, laboratory, and imaging parameters .^10–16^ Yet these models frequently lack sensitivity, generalizability, and external validation that limit their utility in real-world clinical settings. It is well-established that cerebrovascular morphology, including vessel tortuosity, caliber, branching complexity, and collateral development influences infarct progression, reperfusion dynamics, and susceptibility to secondary injury^17,18^. Hence, integrating these cerebrovascular features may substantially enhance risk prediction after EVT ^19^. ML-driven vascular analysis provides an avenue for rapid, accurate, and objective characterization of this anatomical substrate. Our prior pilot work demonstrated that incorporating cerebrovascular morphometry improves the prediction of long-term functional outcomes when combined with conventional variables ^20^. Nonetheless, the existing models for early neurological deterioration and complications largely ignore patient-specific cerebrovascular anatomy and collateral architecture.

In this study, we leverage a validated ML pipeline for automated cerebrovascular segmentation ^21^ and feature extraction from baseline vessel imaging to build predictive models for END and major complications after EVT, including sICH, MBE requiring DHC, and need for tracheostomy/gastrostomy (TC/PEG). We hypothesize that incorporating quantitative vascular morphology and collateral metrics into clinical ML models will significantly improve early complication and outcome prediction compared to conventional approaches, enabling a more individualized and physiologically informed framework for post-EVT care.

## 2. METHODS

### 2.1 Study Design, Population, and Data Acquisition

We retrospectively analyzed data from a prospectively collected cohort of 727 anonymized AIS patients treated with EVT at a comprehensive stroke center between 2019 and 2023. All data use and analysis procedures were approved by the Institutional Review Boards at Inova Fairfax Medical Campus (protocol# INOVA-2024-103) and the University of California, Riverside (Protocol #30175). Deidentified clinical and imaging data were shared in accordance with the “Data Transfer and Use Agreement” established between the two institutions. Patients were included if they had an anterior circulation medium or large vessel occlusion, received EVT, and had complete and evaluable baseline head non-contrast CT (NCCT) and CT angiography (CTA). Patients with incomplete imaging data, severe motion- or contrast-related artifact preventing reliable segmentation, and missing functional outcomes were excluded.

#### Clinical and Demographic Variables

The following baseline demographic and clinical characteristics were extracted: demographics (age, sex, race, and body mass index), baseline modified Rankin Scale (mRS), relevant past medical history (hypertension, diabetes mellitus, dyslipidemia, carotid stenosis, coronary artery disease or myocardial infarction, atrial fibrillation, heart failure, renal insufficiency, prior stroke, and smoking status), admission systolic and diastolic blood pressure, and the NIH stroke scale (NIHSS). Patients were also required to have serial NIHSS assessments at 24 hours, 72 hours, and discharge, as well as a 90-day mRS evaluation.

#### Image Variables Acquisition

ASPECTS was assessed and adjudicated on the presentation NCCT. CTA scans were acquired on clinical multidetector CT scanners with the following parameters: FOV 232 × 232 mm, 512 × 512 matrix, 1.25-mm slice thickness with 0.625-mm spacing, and an in-plane resolution of 0.45 × 0.45 mm. CT perfusion (CTP) data was collected when acquired based on AHA guidelines ^22^. The size of ischemic core (rCBF < 30%) and tissue-at-risk (Tmax > 6 seconds) was obtained from CTP maps provided by RAPID-AI ^23^ and Viz.ai^24^.

#### Laboratory Variables

Admission laboratory values included white blood cell count (WBC), neutrophil and lymphocyte absolute counts and ratio, platelet count, as well as low-density lipoprotein, total cholesterol, serum glucose, creatinine, troponin, and sodium levels.

#### Acute Reperfusion Therapies

Treatment-related variables included IV thrombolytic administration, final adjudicated mTICI score after EVT, door-to-recanalization and onset-to-recanalization times, and medications administered in the first 48 hours, including vasoactive agents, sedative infusions, and hyperosmolar therapy.

### 2.2 Imaging Data Analysis and Feature Extraction

We processed CTA images using our validated AI-based cerebrovascular segmentation pipeline, employing a convolutional neural network (CNN) trained on pre-processed angiographic scans ^20^. The algorithm follows a multi-stage workflow to generate high-fidelity vascular representations. Raw scans are first segmented into a 3D binary vascular map, after which a medial-axis thinning procedure is applied to skeletonize the vessel network and extract vessel centerlines ^21^. This enables reliable estimation of vessel diameters and identification of arterial segments and branching points, including vessels approaching the spatial limits of image resolution. Compared with conventional segmentation approaches, the model demonstrated improved robustness to image-acquisition artifacts such as motion, noise, and inflow effects ^21^. Following segmentation and skeletonization, global morphologic features of the intracranial arterial tree were computed for each patient, including tortuosity, vessel diameter, total arterial length, branch count, and fractal complexity. These quantitative cerebrovascular measures formed the basis for deriving a patient-specific estimate of collateral circulation. To compute a quantitative collateral index (qCI), CTA scans were spatially registered to a standardized CT template ^25^, enabling automated separation of the ipsilateral and contralateral hemispheres. Hemispheric vascular features were then compared to estimate the relative degree of ipsilateral collateralization. The qCI framework was validated against the following clinician-assigned 4-point collateral grading scale ^26^: Grade 0, no detectable ipsilateral collateral supply; Grade 1, partial collateral filling (>0% and ≤50% of the contralateral hemisphere); Grade 2, moderate collateral filling (>50% but <100%); and Grade 3, complete collateralization comparable to the contralateral hemisphere. The automated qCI provided a rapid, objective, and reproducible measure of collateral status that was fully derived from CTA-based vascular morphology and can be integrated directly into predictive modeling.

### 2.3 Outcome Definitions

We developed ML models to predict the following early outcomes and complications, incorporating the extracted quantitative cerebrovascular morphology described above to enhance early prediction. The outcomes were dichotomized (0/1) where appropriate.

#### Symptomatic Intracranial Hemorrhage (sICH)

Hemorrhagic transformation of infarcted tissue is a frequent complication after AIS and reperfusion therapy, though only a subset progresses to symptomatic intracranial hemorrhage (sICH), characterized by a parenchymal hematoma with mass effect and neurological worsening. sICH is thought to be driven by reperfusion injury, endothelial disruption, and impaired autoregulation ^15,16^. Although several clinical and imaging-based predictive scores exist, they often rely on numerous or inconsistently available variables, show limited external validation ^8,16,27^ and do not incorporate cerebrovascular architecture despite the role of vessel fragility, tortuosity, and lumen caliber in hemorrhagic risk ^16^. In this study, we defined sICH as a worsening of ≥4 NIHSS points within 36 hours post EVT, as is used commonly in literature ^8,15^.

#### Malignant Brain Edema (MBE) Requiring Decompressive Hemicraniectomy (DHC)

Large hemispheric infarctions occur in approximately 10% of AIS cases, particularly in patients with complete MCA territory involvement ^28,29^. These infarcts can lead to secondary brain injury through progressive cytotoxic and vasogenic edema, resulting in rising intracranial pressure, midline shift, and rapid neurological deterioration. Decompressive hemicraniectomy (DHC) is a lifesaving intervention that mitigates herniation risk and improves survival ^29^. Numerous demographic, clinical, imaging, and laboratory factors — including age, baseline mRS, prior stroke, hypertension, diabetes, initial NIHSS, ASPECTS, infarct core size, collateral status, and leukocytosis—have been associated with risk of MBE. Despite this, early prediction remains challenging due to heterogeneity in stroke physiology and variability in reperfusion response. This cohort was comprised of patients who developed MBE post EVT and underwent DHC.

#### Tracheostomy and Percutaneous Endoscopic Gastrostomy (TC/PEG)

Neurogenic respiratory failure (NRF) and dysphagia are common consequences of large hemispheric strokes and strategically located subcortical infarcts, affecting up to 40% of AIS patients ^30^. Impaired neuromuscular control of respiration or swallowing may necessitate tracheostomy for prolonged ventilatory support ^31^ and PEG placement for nutritional needs ^32^. Approximately 10–20% of severe AIS patients undergo TC/PEG ^31–33^. Early identification of patients at high risk for respiratory failure or dysphagia is essential for optimizing ICU resources, reducing complications, shortening hospital length of stay, and guiding rehabilitation planning ^33^. However, there are currently no reliable models to predict the need for TC/PEG. This cohort was comprised of patients with NRF post EVT and who underwent a Tracheostomy and a PEG procedure.

#### Early Neurological Deterioration (END) and Improvement (ENI)

END was defined as a ≥4-point increase in NIHSS within the first 24–72 hours after AIS. It occurs in 10–40% of patients, depending on population characteristics and treatment protocols. END is strongly associated with early complications such as infarct expansion, re-occlusion, hemorrhagic transformation, and cerebral edema ^34^. Known risk factors include stroke severity, hypertension, diabetes, dyslipidemia, and inadequate reperfusion. END is tightly linked to long-term disability and mortality ^34^. ENI is most commonly defined as a reduction of ≥4 NIHSS points or an absolute NIHSS of 0–1 and is a well-established early marker of favorable long-term recovery and functional outcomes.

### 2.4 Machine Learning (ML) Model Development

In this study, we employed supervised ML methods to predict multiple post-EVT complications and early outcomes described above using a high-dimensional dataset.

#### Data Preprocessing and Feature Engineering

Predictor variables encompassed demographic characteristics, comorbidities, baseline functional status, admission NIHSS and blood pressure, laboratory markers, administered drugs that affect cerebral blood flow and metabolism, acute reperfusion therapies, and a comprehensive suite of quantitative CTA-derived vascular features, including total arterial length, branch count, diameter, tortuosity, fractal dimension, vessel complexity, and the quantitative collateral index (qCI). CTP-derived measures such as estimated ischemic core and penumbra volume were also included, when available. Data preprocessing was implemented using a scikit-learn ColumnTransformer pipeline, consisted of type harmonization and conversion of all numeric predictors, missing-value imputation (using median imputation for numeric fields and most-frequent imputation for categorical fields), one-hot encoding of categorical predictors (e.g., occlusion site, stroke etiology, smoking status), Z-score standardization of continuous variables, and automated feature selection support (using chi-square–based univariate screening to reduce noise, improve stability, and mitigate overfitting).

#### Model Training and Validation Strategy

To obtain robust and unbiased performance estimates, we used an outcome-specific modeling framework with internal cross-validation and external held-out testing. Each clinical endpoint was modeled independently (per-outcome modeling) to account for varying prevalence and distinct predictor profiles. For each outcome, the dataset was partitioned into 80% training and 20% blind test sets using stratification to preserve class distribution. Within the training set, 5-fold stratified cross-validation generated Out-of-fold (OOF) probability estimates for each patient, simulating prospective model performance without overfitting. We used HistGradientBoostingClassifier, a modern gradient-boosted decision tree (GBDT) algorithm optimized for tabular clinical data. Models were trained with balanced class weights, a learning rate of 0.05, and 400 boosting iterations. Finally, to improve clinical interpretability, each model was wrapped in sigmoid (Platt) calibration, yielding calibrated predicted probabilities for both OOF and test sets. This training pipeline produced two probability vectors per outcome, including the OOF probabilities for internal validation and the blind test probabilities for external validation on unseen patients.

#### Model Comparison with and without Morphology Metrics

To quantify the additive predictive value of cerebrovascular morphology, we trained two models per outcome. A baseline clinical (BC) model without any CTA-derived morphology features and a morphology informed (MI) model that included all clinical, laboratory, treatment, and perfusion variables similar to the BC model plus CTA-derived morphology features (tortuosity, total length, branch count, fractality, and qCI). For each outcome, ROC curves from the two models were overlaid to compare discrimination performance. Differences in AUROC were formally assessed using DeLong testing and metrics such as Net Reclassification Improvement (NRI).

#### Model Evaluation

Models were evaluated using both OOF validation set and blind test predictions according to study discrimination (ROC curves), calibration (Brier score and reliability curves), classification performance (accuracy at 0.5 threshold and Youden’s J optimal threshold), confusion matrices for both thresholds and for both datasets, and risk stratification (patients were categorized into low <0.2, moderate 0.2–0.5, and high >0.5 predicted risk bands). The observed complication rates and cohort counts within these bins were plotted for each outcome. The permutation importance was computed for selected outcomes to quantify the contribution of individual predictors, including vascular morphology, to model predictions.

#### Multitask Patient-Level Risk Profiling

To provide comprehensive personalized risk estimates, we constructed a multitask prediction table for all patients by assembling the calibrated probability outputs for each outcome into a unified matrix. Each patient, therefore, received a vector of predicted probabilities for all complications and early outcomes. The cross-outcome risks were visualized using clustered risk heatmaps, grouping patients by shared risk signatures, radar plots of the distribution of complication risks per outcome, and patient-specific risk profiles supporting individualized clinical interpretation. This framework provided a multidimensional view of patient risk and enables integration of morphology-driven prediction into clinical decision-making.

## 3. RESULTS

A total of 727 AIS patients treated with EVT met the inclusion criteria. After filtering for outcomes with available binary labels (END, sICH, MBE/DHC, TC/PEG), 525 patients remained for model development. The cohort’s mean age was (70±15) years, 48% were female, and the baseline median NIHSS was 16. ENI was seen in 30–35%, and 20-25% developed END. Rates of major complications included sICH in 9%, MBE requiring DHC in 3–5%, and TC/PEG in 12–15% (reported prevalence ranges reflect minor variability across outcome-specific cohorts and train/test splits arising from stratified modeling and missing-label exclusion). At 90 days, over 40% had a poor functional outcome (mRS 3-6) consistent with current trends ^3^.

CTA-derived cerebrovascular morphology variables (total vessel length, branch count, tortuosity, diameter, fractality, and qCI) were extracted for all patients. Figure 1 shows the distribution of the morphological features across groups for the complications. Patients who developed sICH, MBE/DHC, or underwent TC/PEG consistently demonstrated increased vessel tortuosity, reduced vessel diameter, and lower collateral indices (qCI), reflecting impaired microvascular resilience and collateral support.

**Figure 1.**
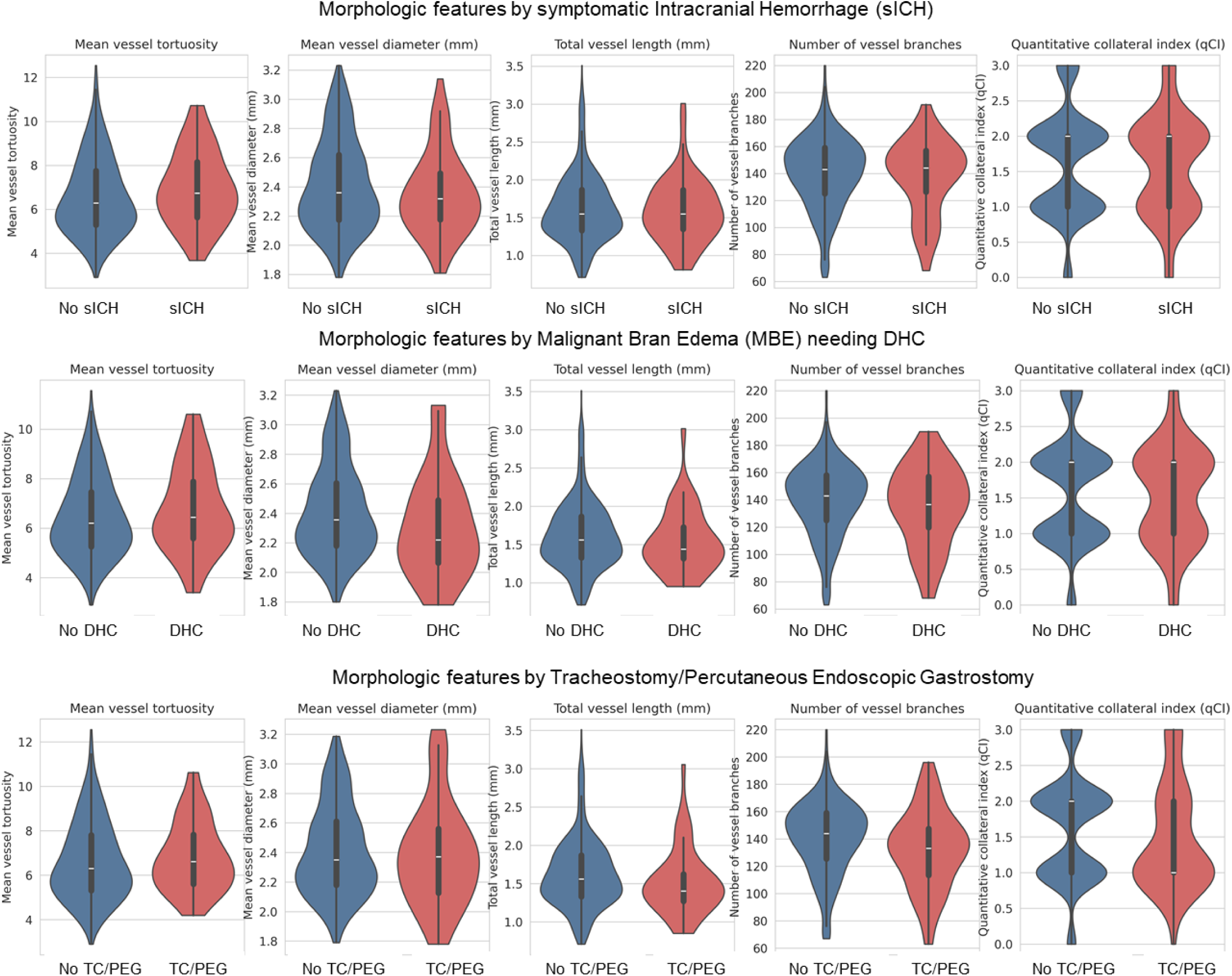
Violin plots comparing quantitative cerebrovascular morphology features between patients with and without mechanistically relevant post-EVT complications that are tied to physiological vascular mechanisms. Each violin represents the full distribution of a given feature, with width proportional to data density, the central marker indicating the median, and the embedded box showing the interquartile range. Comparisons highlight systematic shifts in vascular architecture associated with adverse outcomes.

Figure 2 shows the multi-panel ROC plots comparing the two – basic clinical (BC) and morphology informed (MI) models for the OOF validation and blind test cohorts. Across all modeled endpoints, incorporation of quantitative cerebrovascular morphology consistently improved predictive performance compared to clinical-only models. For each outcome, the MI model demonstrated higher discrimination on both OOF validation and the held-out test set. An improvement was also observed for ENI and dichotomized 90-day mRS (see Supplementary Materials). Importantly, the most pronounced improvement was observed for 90-day mRS prediction, where the MI model demonstrated markedly greater discrimination and calibration compared to the baseline clinical model.

**Figure 2.**
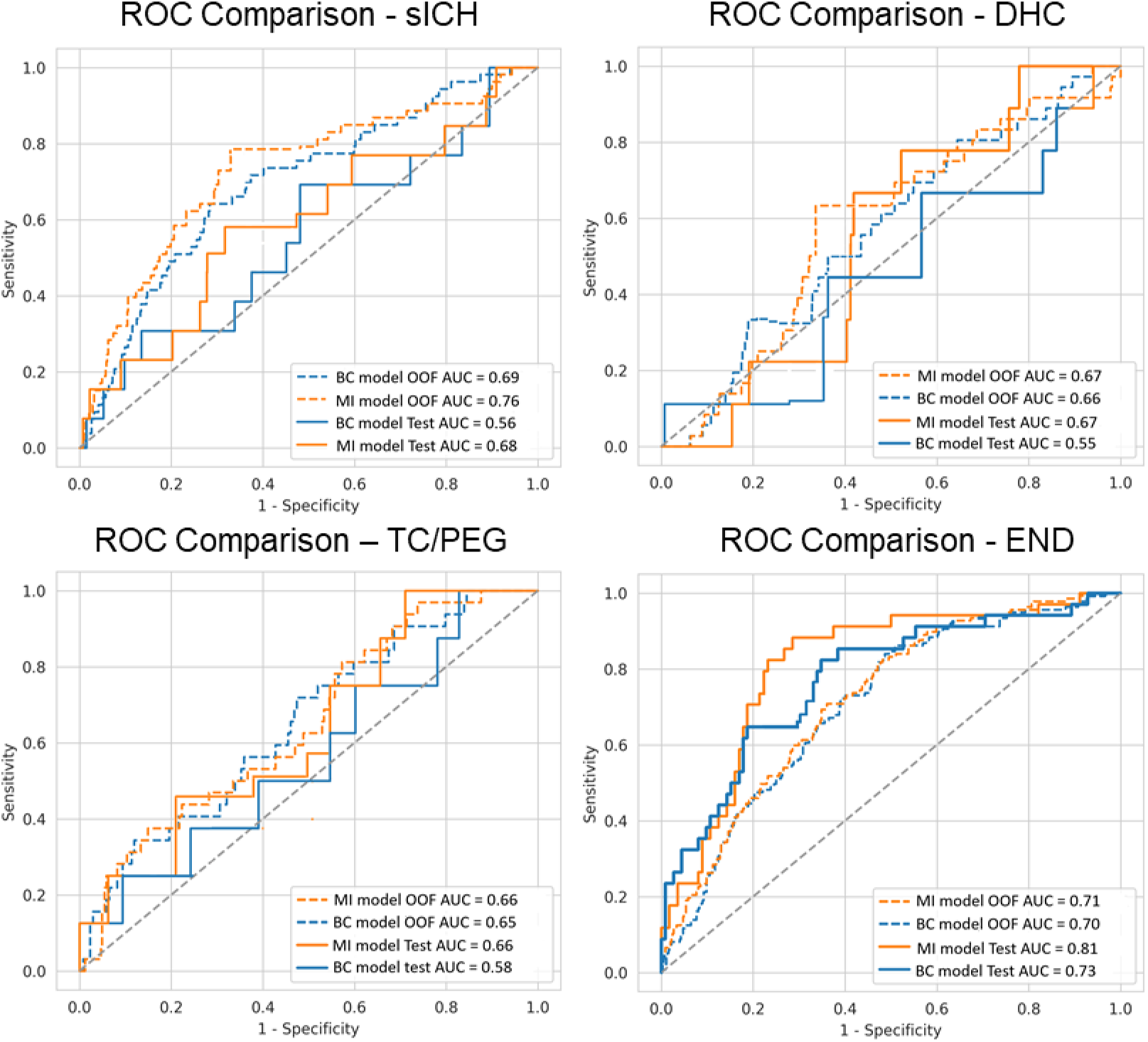
Receiver operating characteristic (ROC) curves comparing the basic clinical (BC) and morphology-integrated (MI) models for post-EVT complication and early outcomes. Each panel displays out-of-fold (OOF) internal hold-out validation and independent blind test-set performance, with the MI model consistently demonstrating improved discriminative ability across multiple endpoints. DHC, decompressive hemicraniectomy; END, early neurological deterioration; MBE, malignant brain edema; sICH, symptomatic intracranial hemorrhage (sICH). AUC values are shown within each panel.

The MI model significantly improved END prediction, achieving AUROC of 0.81 (vs 0.73 for the BC model). The MI model also achieved superior AUROC values relative to the BC model (0.76 versus 0.69 for OOF validation and 0.68 versus 0.56 for test data), for sICH prediction, reflecting enhanced detection of patients at risk for reperfusion-related hemorrhagic injury. Risk-band analysis for sICH occurrence showed that the MI model had improved calibration, higher precision in the high-risk range, and stronger risk stratification. For MBE/DHC prediction, discrimination was modest due to low event count but still improved with adding vascular morphology (0.67 versus 0.56; ΔAUROC was positive with trending significance). In regard to predicting the need for TC/PEG, the MI model also improved prediction, outperforming the BC model by increasing separation between true positive and false positive cases (AUROC 0.66 versus 0.58) and reducing misclassification in the moderate-risk band. Risk stratification showed clear separation of event rates across bands. An improvement was also observed for dichotomized 90-day mRS prediction as well as for predicting early neurological improvement (ENI), where the morphology-enhanced model demonstrated greater discrimination and calibration compared to the baseline clinical model. Since these are not acute complications, the detailed results for these end-points are included in the Supplementary Materials. Together, these findings highlight that the addition of cerebrovascular morphology provides substantial generalizable gains across all major complications and outcome domains, rather than benefiting a single endpoint.

Figure 3 demonstrates the risk stratification results. The patients were classified into low/medium/high risk categories, and the observed event rate per risk level group is seen along with the number of patients. Across all outcomes, the MI models demonstrate clear, monotonic increases in event rates across ascending risk bands. For early neurological deterioration (END) and early neurological improvement (ENI), incorporation of vascular features—including tortuosity, diameter, total vessel length, and qCI—resulted in more accurate identification of patients likely to worsen or improve within 24–72 hours post-EVT. This stratification confirms the models’ ability not only to discriminate high-risk from low-risk patients, but also to provide clinically meaningful, tiered risk groupings that align with actual complication incidence.

**Figure 3.**
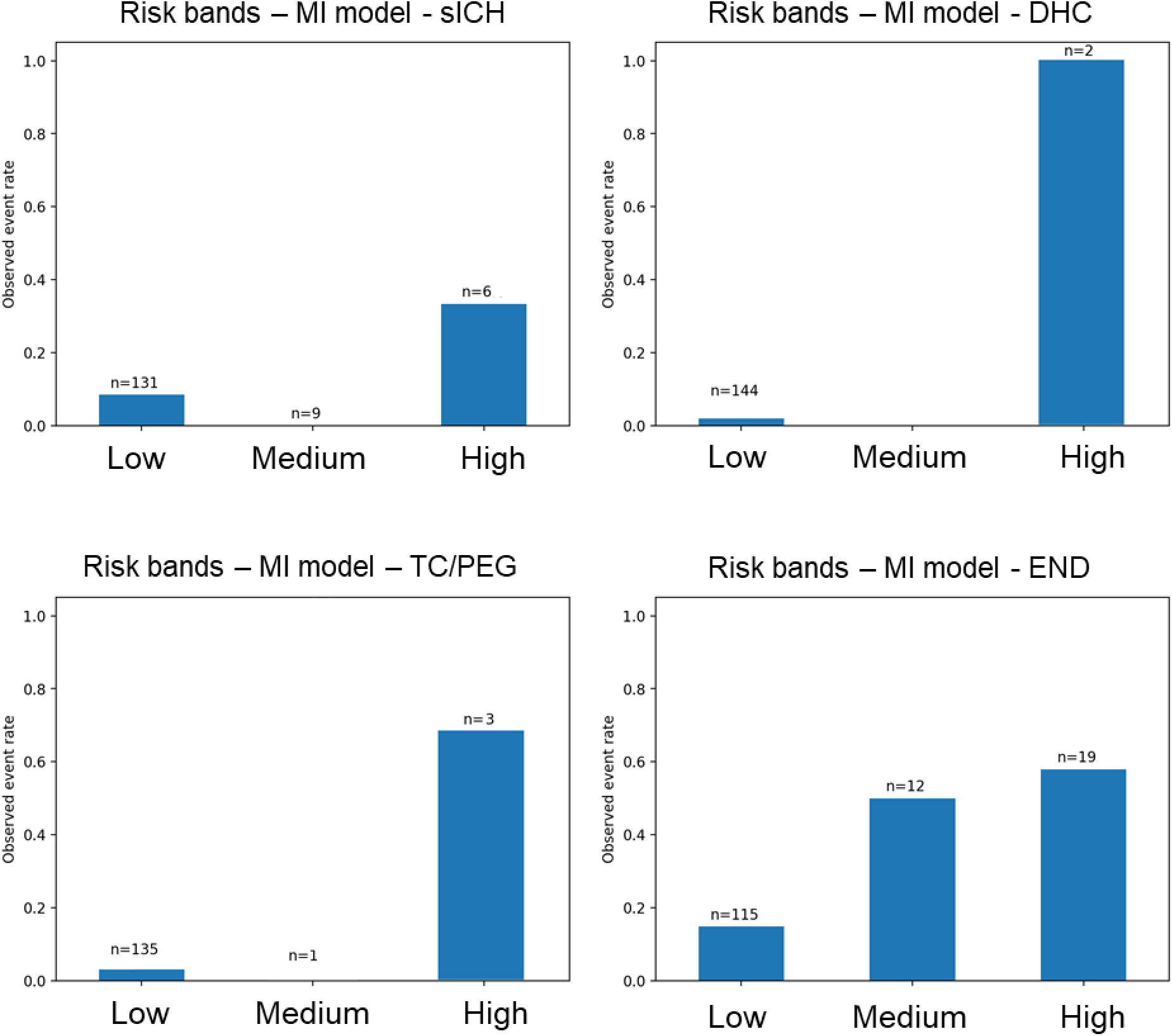
Risk-stratification plots illustrating the relationship between predicted risk probability groups (x-axis) and observed event rate percentages (y-axis) for each modeled post-EVT complication and outcome. Patients were categorized into predefined risk bands—low (<0.20), moderate (0.20–0.50), and high (>0.50)—based on calibrated model-estimated probabilities. Each panel displays the observed event rate (bar height) and corresponding sample size (n-labels) grouped within each risk tier. Symptomatic intracranial hemorrhage, sICH; malignant brain edema requiring decompressive hemicraniectomy, MBE/DHC; tracheostomy/PEG placement, TC/PEG; and early neurological deterioration, END.

Figure 4 shows the permutation feature-importance plots, ranking the top features for each model/outcome based on importance. Across outcomes, quantitative cerebrovascular morphology metrics alongside select clinical and laboratory variables consistently ranked among the most influential predictors. The most informative features included tortuosity (higher in MBE/DHC and sICH), diameter (smaller in high-risk patients), and branch count / total vessel length (smaller in high-risk patients). Fractality and qCI were both higher in low-risk patients, indicative of a protective mechanism. This confirms that vascular architecture encodes biological information reflecting reserve capacity, tissue vulnerability, and susceptibility to secondary injury.

**Figure 4.**
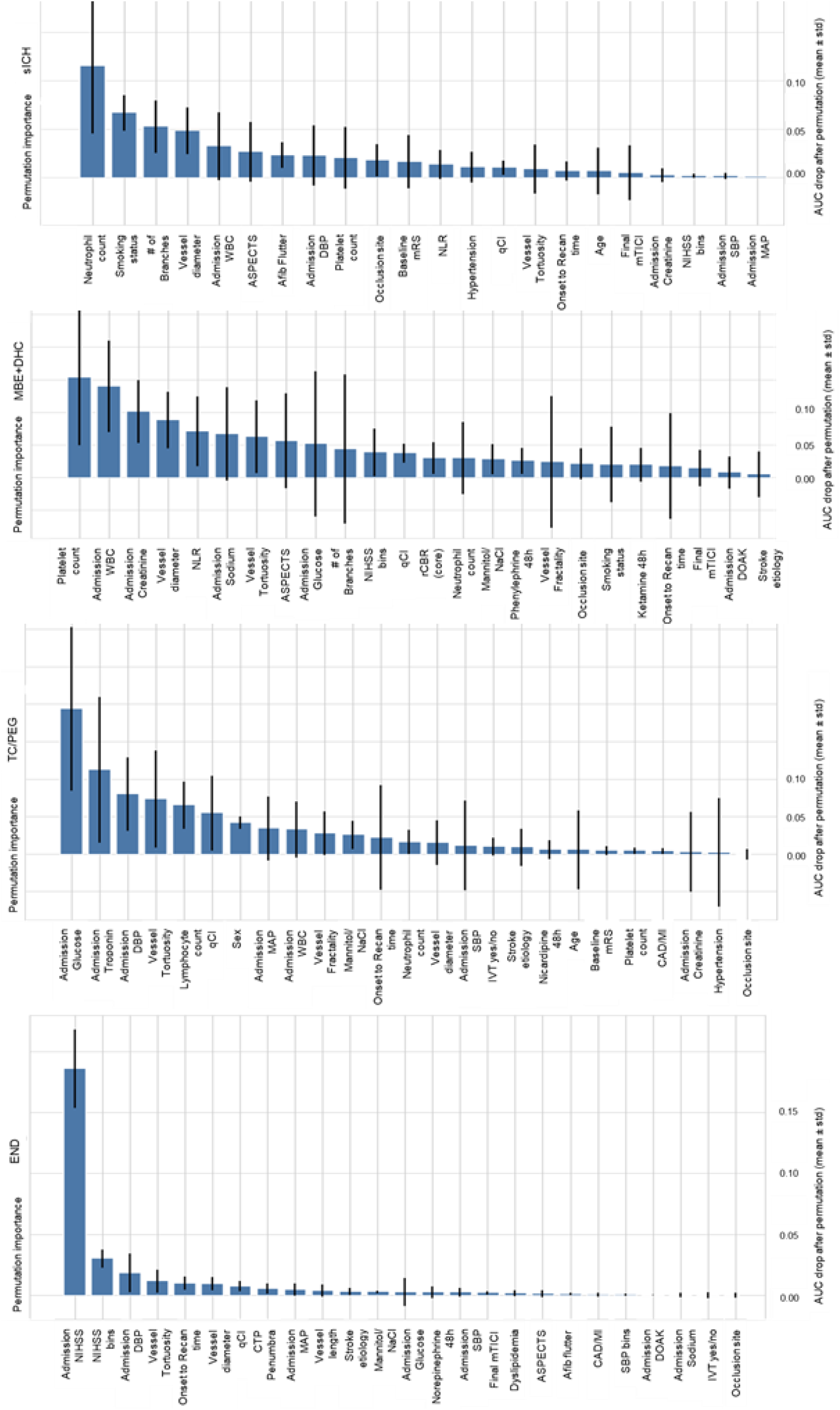
Permutation feature-importance analyses for post-EVT complication and outcome models. Each panel displays the decrease in model performance (ΔAUROC) resulting from random permutation of a single predictor variable, thereby quantifying that predictor’s unique contribution to model discrimination. Higher bars indicate greater degradation in AUROC when the feature is disrupted, reflecting stronger predictive influence.

Overall statistical comparison of the models, using DeLong-style AUROC comparisons, visually highlights these gains for all outcomes when morphology was added (Figure 5), with consistently right-shifted effect sizes. Outcomes with statistically significant improvement included END and sICH with *p*<0.05 (ENI and mRS end-points included in Supplementary Materials). MBE/DHC trended toward improvement but did not reach significance due to low event count. Net Reclassification Improvement (NRI) and Integrated Discrimination Improvement (IDI) further demonstrated enhanced risk discrimination and probability separation, particularly for patients with higher risk of sICH and END (Table 1).

**Figure 5.**
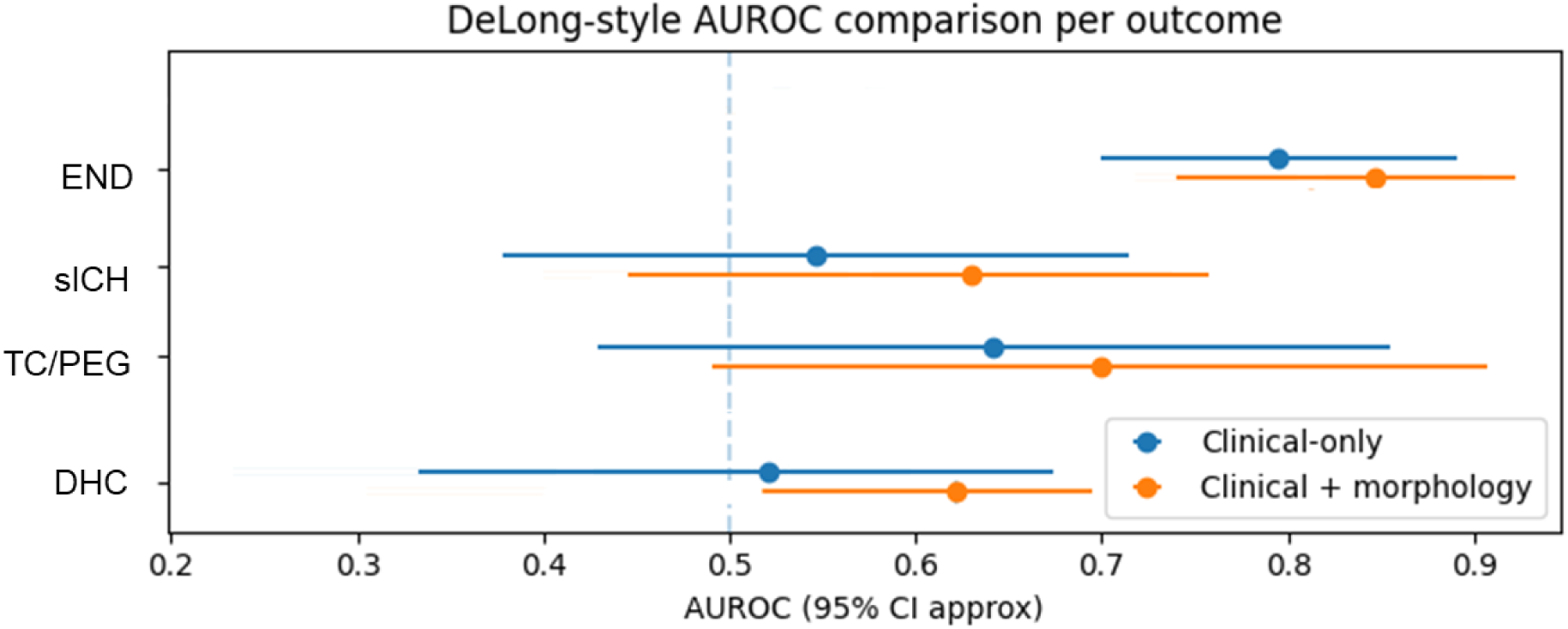
Comparison of model discrimination for each outcome using full (clinical + morphology) versus clinical-only models. AUROC values are displayed side-by-side, with statistical significance of differences assessed using DeLong’s test. Positive AUROC shifts indicate improved discrimination.

**Table 1.**
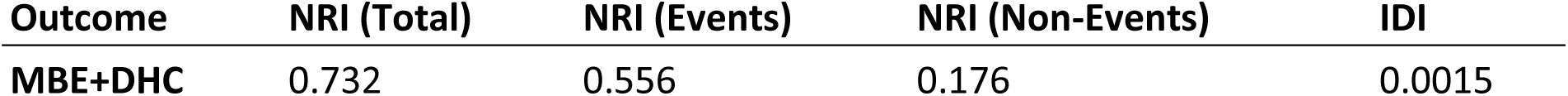
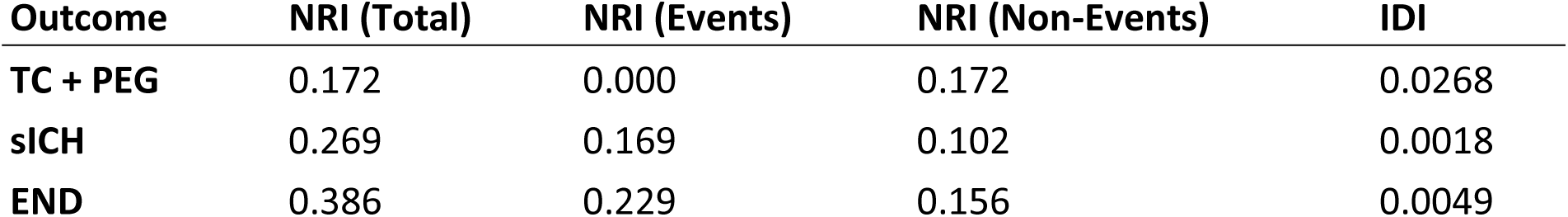
Net Reclassification Improvement (NRI) and Integrated Discrimination Improvement (IDI) comparing the morphology-augmented models to the clinical-only models across all complications and early outcomes. Positive NRI values indicate improved risk stratification, with NRI*events* reflecting better identification of true high-risk patients and NRI*non-events* indicating reduction of false-positive classifications. IDI quantifies improved separation between predicted risks for events vs. non-events. The largest reclassification gains were observed for MBE requiring DHC, sICH and END demonstrating substantial added value of cerebrovascular morphology in prognostic modeling (ENI and mRS in Supplemental Material).

We used a multitask framework to generate individualized risk profiles across all complications. Figure 6 shows patient-level probability maps and radar plots for distinct patient subgroups (identified by risk-heatmap clustering) with coherent multimorbidity patterns. High-risk clusters often had poor collateralization, extensive vascular narrowing, and higher tortuosity. Some patients were selectively high-risk for sICH while others were high-risk for requiring TC/PEG, demonstrating the benefit of individualized prediction rather than one-size-fits-all risk models. These visualizations aim to provide interpretable evidence to guide monitoring levels, resource allocation, and early intervention decisions.

**Figure 6.**
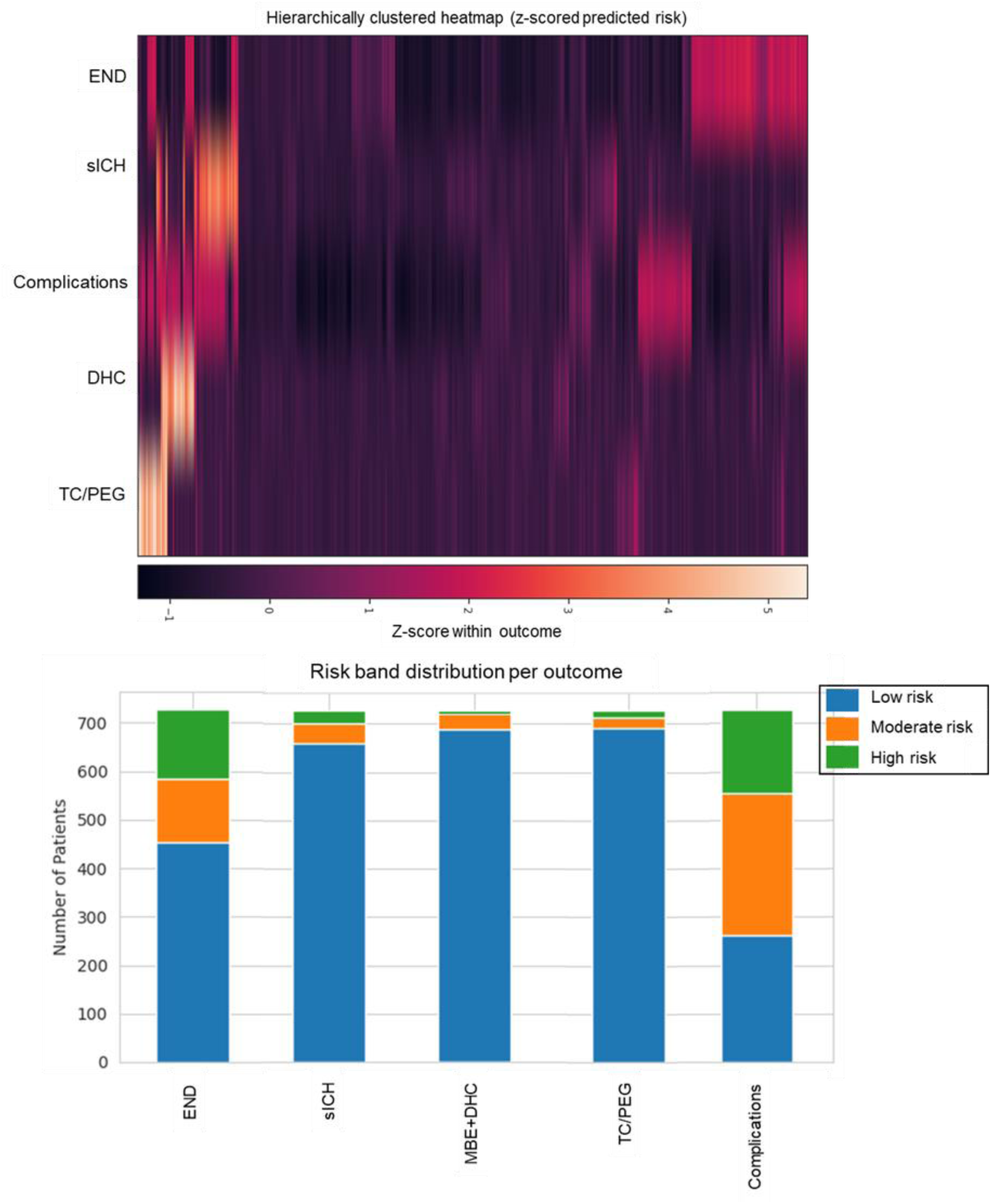
The top panel displays a hierarchical clustered heatmap of predicted probabilities for all complications and early outcomes, revealing coherent patient subgroups with shared high-risk or low-risk vascular–clinical profiles. The bottom panel illustrates outcome-specific risk-band distributions (low <0.2, moderate 0.2–0.5, high >0.5), depicting observed event rates and sample sizes within each band. Together, these visualizations demonstrate the model’s ability to differentiate patient phenotypes and support clinically meaningful stratification for individualized post-EVT risk assessment.

## 4. DISCUSSION

In this study, we demonstrate that incorporating quantitative cerebrovascular morphology and collateral grading metrics into prediction frameworks meaningfully enhances the early identification of post-EVT complications and short-term outcomes in AIS. Across all modeled endpoints, the morphology-informed (MI) models consistently outperformed basic clinical (BC) models in both discrimination and calibration.

Although several clinical and imaging-based predictive scores exist for AIS complications, they often rely on numerous or inconsistently available variables and show limited external validation ^9,35,36^. Therefore, early prediction remains challenging due to heterogeneity in stroke physiology and variability in reperfusion response, leading to low specificity (AUROC values typically between 0.55 – 0.71 and specificity 0.51 - 0.58) ^10,12,16,16,36^. These scores do not incorporate cerebrovascular architecture despite its known role in brain tissue response to ischemia and reperfusion and likely contribution to early major complications ^16^. Traditional prognostic models rely heavily on demographics, presentation NIHSS and ASPECTS scores, and treatment parameters. Prior ML models to predict complications also mostly rely on clinical or perfusion-derived metrics. Few incorporate vascular morphology, and none to our knowledge provide a comprehensive per-patient probabilistic risk matrix across multiple interdependent complications. Moreover, current scoring systems such as HIAT2, SEDAN, DRAGON, or EDEMA scores ^27,35,37–39^ rely on manually extracted variables and lack external validation, generalizability, or integration of vascular structure. The consistent improvements observed in our MI models suggest that incorporating patient-specific vascular architecture and collateral status provides unique and complimentary predictive information that augments traditional clinical and imaging covariates.

Quantitative vascular morphology metrics reflected underlying cerebrovascular structural vulnerability and reduced collateral reserve—features not typically captured by conventional clinical or perfusion metrics—thereby aiding in the early identification of patients at increased risk for post-EVT deterioration. DeLong testing further confirmed that these differences were significant for a majority of outcomes. Integrated discrimination improvement (IDI) analysis supported this by showing increased separation between predicted risks in event vs. non-event patients, indicating that the MI model assigns appropriately higher probabilities to true complication cases while slightly reducing risk overestimation in non-events. Complementarily, the Net Reclassification Improvement (NRI) analysis showed that adding morphology metrics systematically reclassified more susceptible patients into more clinically appropriate higher risk categories, reducing under-estimations seen often in clinical-only models. Collectively, these metrics underscore that vascular morphology introduces a qualitatively different predictive dimension, refining both individualized risk estimation and population-level stratification.

### 4.1 Biological Interpretation of Morphologic Predictors

Permutation importance analyses revealed that morphological features were strong contributors to prediction. These findings align well with known pathophysiological mechanisms and reinforce the central biological premise of our work that cerebrovascular morphology features are mechanistically integral determinants of downstream reperfusion injury and early neurological trajectories ^7,19,40,41^. For instance, qCI, a surrogate for hemispheric collateral filling, was among the most informative predictors across early outcomes, reinforcing the critical protective role of collaterals in mitigating the ischemic cascade. Weak or underdeveloped collaterals as well as chronic hypertension with arteriosclerosis can lead to reperfusion injury, incomplete microvascular reperfusion, and persistent metabolic stress that in turn increase the risk of hemorrhagic transformation and edema formation.

The well-established risk-factors for sICH that have been commonly used for predictive modeling are age, NIHSS, blood glucose level, platelet count, hyperdense artery signs on NCCT, and elevated systolic blood pressure ^8,15^. Vascular abnormalities such as heightened tortuosity, vascular rarefaction, and irregular/narrowed lumen can be linked to vessel wall weakening and endothelial dysfunction, further increasing the chances of sICH manifestation or edema formation ^19^. Along with the well-known risk factors, we saw that elevated neutrophils, smoking status, and higher diastolic blood pressure emerged as key predictors of sICH (Figure 5), reflecting the combined effects of systemic inflammation, endothelial dysfunction, and increased vascular stress that promote blood–brain barrier breakdown and hemorrhagic transformation after reperfusion.

In contrast, MBE requiring DHC was more strongly associated with markers of inflammatory burden and metabolic stress, including elevated platelets, WBC count, and neutrophil to lymphocyte ratio, as well as serum creatinine and blood glucose which reflect heightened immune activation, microvascular injury, impaired tissue clearance, and greater susceptibility to vasogenic edema. Collectively, these biologically complementary features highlight interaction between various physiological pathways that lead to worsening microvascular integrity, thereby predisposing patients to distinct yet related post-EVT complications.

Our study advances the field by introducing quantitative cerebrovascular geometry as a robust imaging biomarker, using rigorously validated pipelines with stratified cross-validation, calibrated probabilities, and independent hold-out testing, demonstrating superiority through formal statistical comparison and delivering clinically interpretable risk tools, including patient-level risk matrices, and clustered risk heatmaps.

### 4.2 Clinical Utility: Precision Risk Stratification and Patient-Level Risk Profiles

A major strength of this study is the generation of patient-level calibrated risk profiles, where each patient receives a probability estimate for the occurrence of each major complication. Risk-band visualizations further showed clear separation between low-, moderate-, and high-risk groups, providing a natural framework for clinical decision support. Importantly, the morphology-augmented model sharpened the boundaries between these strata, reducing indeterminate (“gray zone”) classifications. Such stratification is especially impactful in EVT populations, where small differences in patient trajectory translate to dramatically different resource needs and clinical outcomes. This multidimensional risk matrix aims to benefit clinically in multiple ways, such as improving early triage and resource deployment for the patients at elevated risk for impending MBE or sICH and enhanced communication with families, supported by concrete, individualized risk estimates rather than broad population averages.

### 4.3 Strengths and Limitations

Key strengths of this study include using a prospectively collected large real-world EVT cohort, harmonized preprocessing, and a fully automated imaging pipeline that reduces variability and manual bias. The models were built using modern gradient-boosting frameworks, optimized for tabular clinical data, and underwent stringent validation.

Our study also has limitations. The single-center dataset may constrain generalizability. External multicenter validation is needed, particularly for low-incidence outcomes such as DHC. Additionally, although our automated segmentation model has been validated, imaging artifacts, segmentation errors, or severe hypoperfusion may still affect feature extraction. The AUROC values for most models are modest due to the low incidence of complications in the overall data and even more so in the test datasets. This could be mitigated with a larger dataset with a higher number of patients who were treated for complications, although that approach runs the risk of the data not being truly representative of the actual incidence rates in the population.

### 4.4 Conclusions and Future Directions

Our study advances the field by introducing quantitative cerebrovascular geometry as a robust imaging biomarker, using rigorously validated pipelines with stratified cross-validation, calibrated probabilities, and independent hold-out testing. Future work will include multicenter external validation and integration of NCCT and CTP texture features. Deep learning–based multitask architectures could further enhance joint modeling of correlated outcomes. Ultimately, embedding vascular morphology into routine stroke workflows has the potential to fundamentally shift EVT aftercare from reactive to anticipatory, and improve safety, efficiency, and long-term recovery.

## Data Availability

Data was acquired under an IRB approval at the University of California and is not public but can be shared upon request to the seniour author.

## Acknowledgements and Funding

This study was supported by the National Institutes of Health (NIH), the National Institute of Neurological Disorders and Stroke (NINDS) (Grant Numbers: R01-NS131554, R03NS108167) and Inova SEED grants GRT1000000237, GRT1000000288.

